# Exploring the Influence of Lifestyle, Social Health, and Demographic Factors on Psychological Well-being and Engagement Levels

**DOI:** 10.64898/2026.02.09.26345884

**Authors:** Niranjana Arun Menon, Md Rafiqul Islam, Mohamed Reda Bouadjenek, Shoaib Jameel, Eran Segal, Imran Razzak

## Abstract

Loneliness and psychological well-being are increasingly recognized as critical public health concerns, yet their multi factorial determinants remain poorly understood. Traditional research often examines demographic, lifestyle, or social variables in isolation, yielding fragmented insights that overlook complex psychosocial interactions. In this study, we leverage a rich behavioral and psychological dataset from the Human Phenotype Project (HPP) to examine how lifestyle behaviors, social health indicators, and demographic characteristics collectively influence mental health outcomes. Employing advanced machine learning (ML) methods, including feature engineered representations, classical predictive models, and Large Language Model (LLM) classifiers, we identify latent psychosocial patterns associated with loneliness and psychological symptoms. Our approach combines predictive performance with interpretability, enabling the identification of key drivers of well-being across heterogeneous populations. Results indicate that certain lifestyle and social engagement factors consistently correlate with lower loneliness and improved psychological health, while other influences are context-dependent. This work demonstrates the potential of integrating computational modeling with psychological theory to reveal complex, multidimensional determinants of mental health, offering insights for targeted interventions, digital health applications, and evidence-based public health strategies.

## 1 Introduction

Loneliness and psychological well-being are increasingly being recognized as critical public health concerns, with profound implications for both individual functioning and societal resilience. Traditional research has typically examined these outcomes through isolated demographic, behavioral, or social variables, producing fragmented insights that overlook the complexity of real-world psychosocial experiences [6, 25, 26]. Yet emerging evidence, including the influential work of Holt-Lunstad et al. [7] suggests that loneliness and psychological symptoms arise from dynamic, interconnected patterns shaped by a constellation of demographic, lifestyle, and social health factors.

Advances in computational social science and machine learning enable modeling of complex human experiences, including feature engineering [15] and LLM-based trait inference [20]. However, their application to mental health research—especially for interpretable and ethical insights—remains limited, despite their potential to inform digital health interventions and public health strategies. In this study, we integrate psychological theories with data-driven modeling to better characterize the multidimensional nature of loneliness and psychological health. As illustrated in Figure 1, we aim to move beyond single-factor analyses by examining how demographic characteristics, lifestyle behaviors, and social health indicators interact to shape psychological outcomes. This effort is grounded in two overarching research questions:

**Figure 1:**
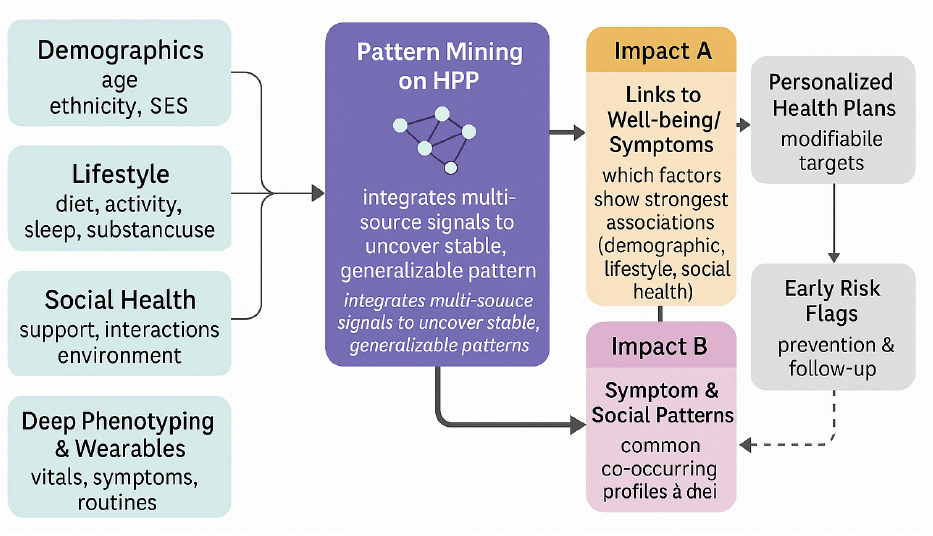
Conceptual framework linking multi factorial determinants to loneliness outcomes. This framework illustrates a theory-driven approach to modeling loneliness and guides the subsequent research questions.

- **RQ1:** Which lifestyle, social health, and demographic factors are associated with higher psychological well-being or psychological symptoms?
- **RQ2:** What recurring configurations of psychological and social health factors are associated with different loneliness outcomes, and how do these configurations relate to loneliness status?

To address these questions, we leverage the Human Phenotype Project (HPP) dataset [18], a rich repository of sociodemographic, behavioral, and psychological assessments. By using advanced machine learning methods such as target encoding, feature engineered representations, classical predictive models, and LLM-based classifiers, we evaluate both predictive performance and interpretability of latent psychosocial patterns. This combination of methodological rigor and social relevance positions our work at the intersection of mental health research and responsible AI. By uncovering the multi factorial determinants of loneliness and psychological well-being, our findings offer insights into how computational methods can enhance understanding of complex psychosocial phenomena and support human-centered digital health technologies. Key contributions include:

- Demonstrating that lifestyle, social health, and demographic factors jointly influence psychological outcomes, highlighting the multi factorial nature of loneliness.
- Implementing a machine learning pipeline combining feature engineering, classical models, and LLM-based classifiers to identify key predictors and latent patterns in social-health data.
- Performing feature contribution analyses to reveal the most influential variables, supporting interpretable, data-driven insights for digital mental health interventions.

Technical contributions include:

- Developed a unified preprocessing pipeline integrating 108 sociodemographic, behavioural, and psychological variables across three HPP datasets (5,944 participants).
- Compared categorical encoding strategies, showing target encoding preserves class structure better than label encoding in latent-space analyses.
- Applied Recursive Feature Elimination (RFE) to select 15 key predictors, improving minority-class detection and interpretability.
- Conducted univariate logistic-regression augmentation to quantify each feature’s contribution to loneliness prediction. Benchmarked classical classifiers such as logistic regression, tree ensembles, and boosted models across feature and encoding configurations.
- Evaluated a compact reasoning-capable large language model with parameter-efficient fine-tuning and chain-of-thought prompting.

## 2 Related Work

In this section, we highlight evidence linking lifestyle behaviours, social health, and demographic factors to psychological well-being and engagement. By examining previous work that has employed both traditional and data-driven methodologies, we aim to justify the relevance of these factors and the approaches chosen to investigate their effects.

### 2.1 Lifestyle Determinants

Researchers have linked lifestyle behaviors to mental health outcomes, though effect sizes vary. Nutritional interventions show modest improvements in depressive symptoms from diets rich in omega-3 and vitamins, but heterogeneity and publication bias persist [1, 4, 10, 14]. In contrast, physical activity and sleep quality show more consistent associations with mood, where higher habitual activity and better sleep predict lower depressive symptoms, while disrupted sleep is a strong risk factor for depression and anxiety [21]. Substance use (alcohol, tobacco, recreational drugs) further exacerbates mental-health risk and confounds associations with other lifestyle factors, necessitating careful covariate control. As studies increasingly move beyond single-behaviour analyses, behavioural clustering approaches identify person-level lifestyle patterns, typically “healthy”, “mixed”, and “unhealthy”, that demonstrate stronger links to mental health than individual behaviours [11]. Integrating individual lifestyle measures with cluster membership thus provides a more comprehensive framework for understanding how demographic, behavioural, and social-health factors collectively shape psychological well-being.

### 2.2 Social Health and Loneliness

Social connectedness and loneliness are among the strongest predictors of psychological distress. Loneliness is typically measured using the UCLA Loneliness Scale [2], though invariance across groups requires verification. Intervention studies and systematic reviews show that loneliness is modifiable, with psychological, social-skills, community-based, and digital interventions producing small to moderate improvements depending on population and baseline severity [22], though heterogeneity and limited long-term follow-up remain challenges. Meta-analytic findings by Lasgaard et al. [9] similarly report benefits of cognitive–behavioural and social-skills programs for older adults and students but emphasise inconsistent effects and unclear mechanisms. Recent umbrella and rapid reviews further conclude that while loneliness can be reduced, more rigorous, mechanism-driven trials are needed. For RQ2, these insights underscore the importance of assessing loneliness using validated scales and modelling it both as an outcome (capturing how symptom or social-health classes differ in loneliness) and as a predictor or mediator (examining how loneliness contributes to psychological symptoms and engagement), reflecting its central role in shaping mental-health trajectories.

### 2.3 Patterns of psychological and social-health symptoms

Recent work has moved beyond isolated factor analysis to examine psychological symptoms as interacting components within network-based frameworks. Rather than attributing outcomes to latent constructs alone, these approaches present symptoms as dynamically connected systems. For example, Pan et al.[16] showed that symptoms such as sleep disturbance, stress, and irregular emotions can form interlinked modules that influence overall mental health stability, while Chuinsiri et al.[3] used unsupervised learning to identify symptom clusters associated with socioeconomic disadvantage and functional impairment. Together, these studies demonstrate how network and clustering methods can uncover latent symptom structures and their social correlates, aligning with RQ2’s focus on symptom interrelationships and loneliness.

Following the literature, we present examples based on the HPP variables structure (discussed more in section 3.1) to demonstrate how such interaction patterns may appear in large-scale observational data. For example, higher loneliness and stress can be linked with reduced daily physical activity, shorter sleep, and lower social engagement, while a contrasting pattern involves stronger social connections and better sleep co-occurring with higher daily activity and leisure participation. Alternatively, higher social engagement may coincide with more consistent physical activity and lower reported psychological distress, forming a mutually reinforcing cluster. These examples illustrate how HPP variables can form cohesive interaction patterns rather than isolated symptoms, consistent with contemporary network-based and clustering approaches.

### 2.4 Shaping the Research Questions

The reviewed literature highlights the complex and interdependent nature of mental health determinants, spanning behavioural, social, and network-level processes. Lifestyle factors such as nutrition, physical activity, sleep, and substance use interact with social connectedness and loneliness to shape mental health trajectories in multifaceted ways. Recent computational frameworks, including clustering and network-based approaches, further suggest that psychological well-being arises from dynamic interactions among symptom modules rather than isolated predictors. Collectively, these findings motivate integrative modeling strategies that capture behavioural, social, and structural interrelations.

In contrast to unsupervised or network-centric approaches, this study adopts a supervised, interpretability-oriented framework that directly links observable psychological, social, and lifestyle attributes to loneliness outcomes. We examine how combinations of measurable factors relate to loneliness status and quantify their relative contributions, rather than merely inferring latent symptom structures. This design prioritises interpretability and scalability, supporting large-scale screening and intervention-oriented applications.

## 3 Methodology

This section describes the datasets, preprocessing workflow, encoding strategies, feature selection procedures, and modelling frameworks used to develop loneliness prediction models within the HPP cohort.

### 3.1 Dataset Preprocessing

The Human Phenotype Project (HPP) dataset[18] is a large-scale collection of human behavioral, lifestyle, social, and demographic information designed to study how everyday factors influence psychological well-being. It integrates measures such as sleep habits, physical activity, nutrition, social connections, socioeconomic status, and self-reported mental health indicators. The dataset enables researchers to explore how lifestyle patterns, social health, and demographic characteristics interact to shape emotional functioning, stress levels, and overall psychological resilience.

In this study, we used three population-scale sub-datasets, namely `053: Sociodemographics, 055: Lifestyle` and `Environment`, and `057: Psychological and Social Health`. After harmonizing these datasets using the HPP-defined unique participant identifiers, the final analytical cohort consisted of 5,944 unique individuals and a combined feature space of 108 variables. All datasets were linked via pseudonymised participant identifiers compliant with transparency and data protection regulations; no personal or identifying information was accessed. Numerical variables and single-categorical attributes were retained as the primary feature space for supervised learning. Only individuals with complete records across all three datasets were included. The health_mental_current_lonely variable served as the ground-truth target label for all classification experiments, providing the three-category outcome (“Yes”, “No”, “Do not know”) used to define the loneliness classes. The dataset exhibits class imbalance, with 5,023 “Not Lonely” (84.5%), 705 “Lonely” (11.9%), and 216 “Do not know” (3.6%) responses. To mitigate bias, we used class-weighted loss functions in all models. For tree-based models, we set `class_weight=‘balanced’`. No synthetic oversampling (e.g., SMOTE) was applied to avoid artificial feature distortions, but this may contribute to lower recall for minority classes (Section 3.4).

Table 1 presents selected example columns from the dataset to illustrate its activity and demographic structure.

**Table 1:** Excerpt of the merged dataset containing 5,944 participants prior to encoding.

|  | country_of_birth | activity_walking_pace | activity_stairs_daily | activity_walking_pleasure_monthly | activity_walking_pleasure_duration |
| --- | --- | --- | --- | --- | --- |
| 1 | Ukraine | Brisk pace | 1–5 times a day | 2–3 times in last 4 weeks | Over 3 hours |
| 2 | Israel | Brisk pace | 1–5 times a day | Once a week | Between 30 min and 1 hour |
| 4 | nan | Brisk pace | More than 20 times a day | 4–5 times a week | Between 1 and 1.5 hours |
| 6 | Israel | Brisk pace | More than 20 times a day | nan | nan |
| 8 | Israel | Brisk pace | nan | 2–3 times in last 4 weeks | Between 2 and 3 hours |
| ⋮ | ⋮ | ⋮ | ⋮ | ⋮ | ⋮ |
| 10635 | Israel | Steady avg pace | Prefer not to answer | nan | nan |
| 10636 | Israel | Steady avg pace | 11–15 times a day | Once a week | Between 30 min and 1 hour |
| 10638 | Israel | Steady avg pace | More than 20 times a day | nan | nan |
| 10639 | Israel | Steady avg pace | More than 20 times a day | 2–3 times a week | Between 30 min and 1 hour |
| 10641 | Israel | Steady avg pace | More than 20 times a day | 2–3 times a week | Between 1.5 and 2 hours |

### 3.2 Dataset Encoding

The unified HPP dataset was processed through a structured pipeline, including missing value imputation and label encoding. To prevent data leakage, target encoding of categorical features was performed only on the training set after the 80/20 train-validation split, with validation features encoded using target statistics derived from the training data. This stage also included normalization of continuous attributes and construction of the final feature matrix used across all baseline and LLM models. Three encoding strategies: one-hot, label, and target encoding [13] were evaluated for this dataset. One-hot encoding was excluded due to the dataset’s high dimensionality of 108 variables and over 200 categories, which would yield sparse, computationally inefficient representations. Furthermore, based on literature comparing label and target encoding, target encoding was chosen for its ability to handle high-cardinality features, reduce estimator variance, preserve class-conditional structure, and improve empirical performance in tabular learning [5, 17, 24].

To further demonstrate the difference of target and label encoding, we conducted PCA analysis on the target-encoded and label encoded features. The PCA on target encoded features (Figures 2a and 3a) shows separation between classes, however these visualizations do not represent inherent psychosocial patterns and are included only to illustrate the geometric effects of encoding strategies. For true latent structure discovery, we rely on downstream model performance and feature importance analyses (Section 3.4). In contrast, label encoding collapses categorical levels in ways that produce diffuse and poorly aligned clusters. While not definitive, these patterns consistently support the adoption of target encoding for subsequent modeling, motivating its use as a more informative representation for psychosocial classification tasks.

**Figure 2:**
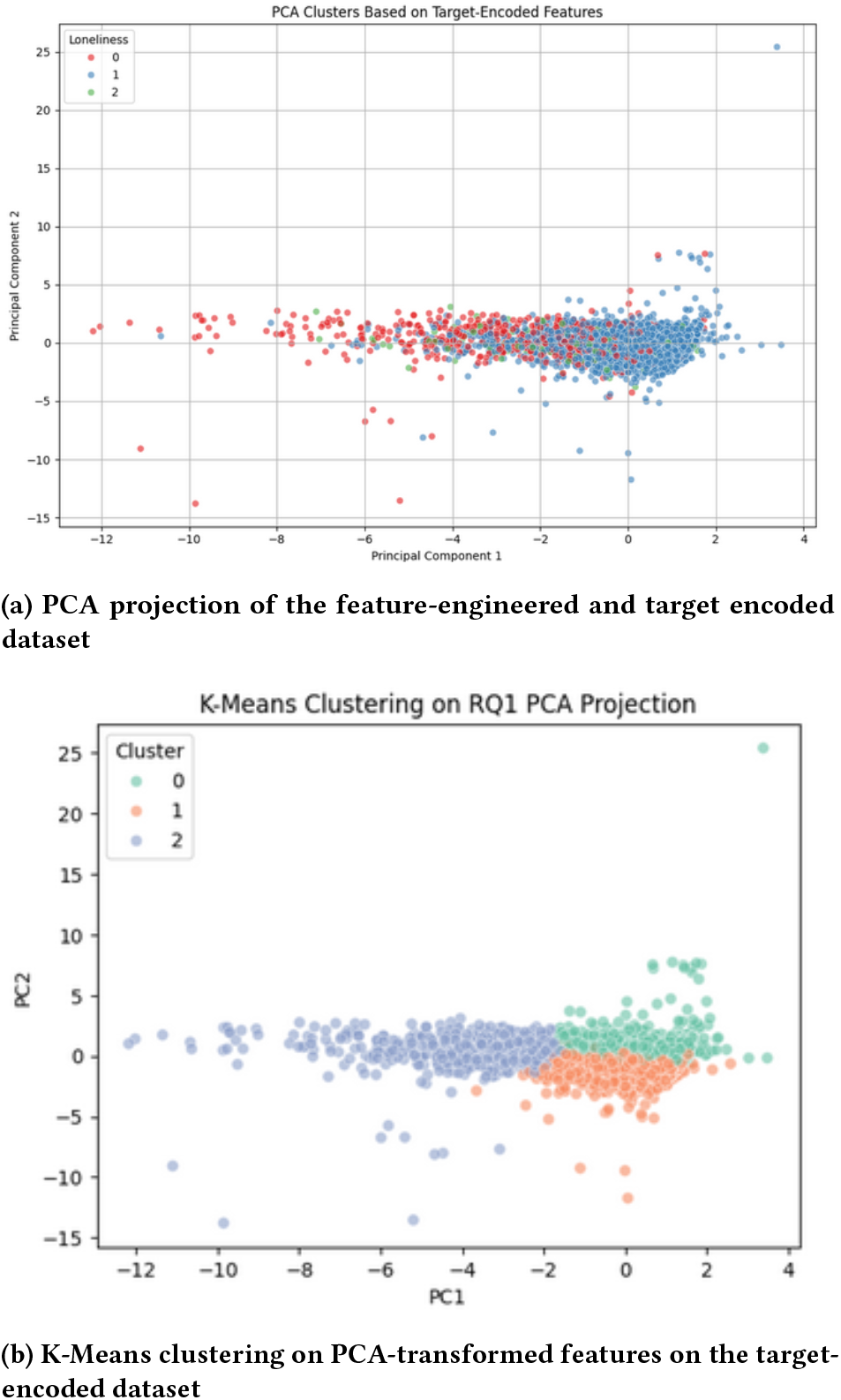
PCA and K-Means visualisations showing latent structure after feature engineering and target encoding.

**Figure 3:**
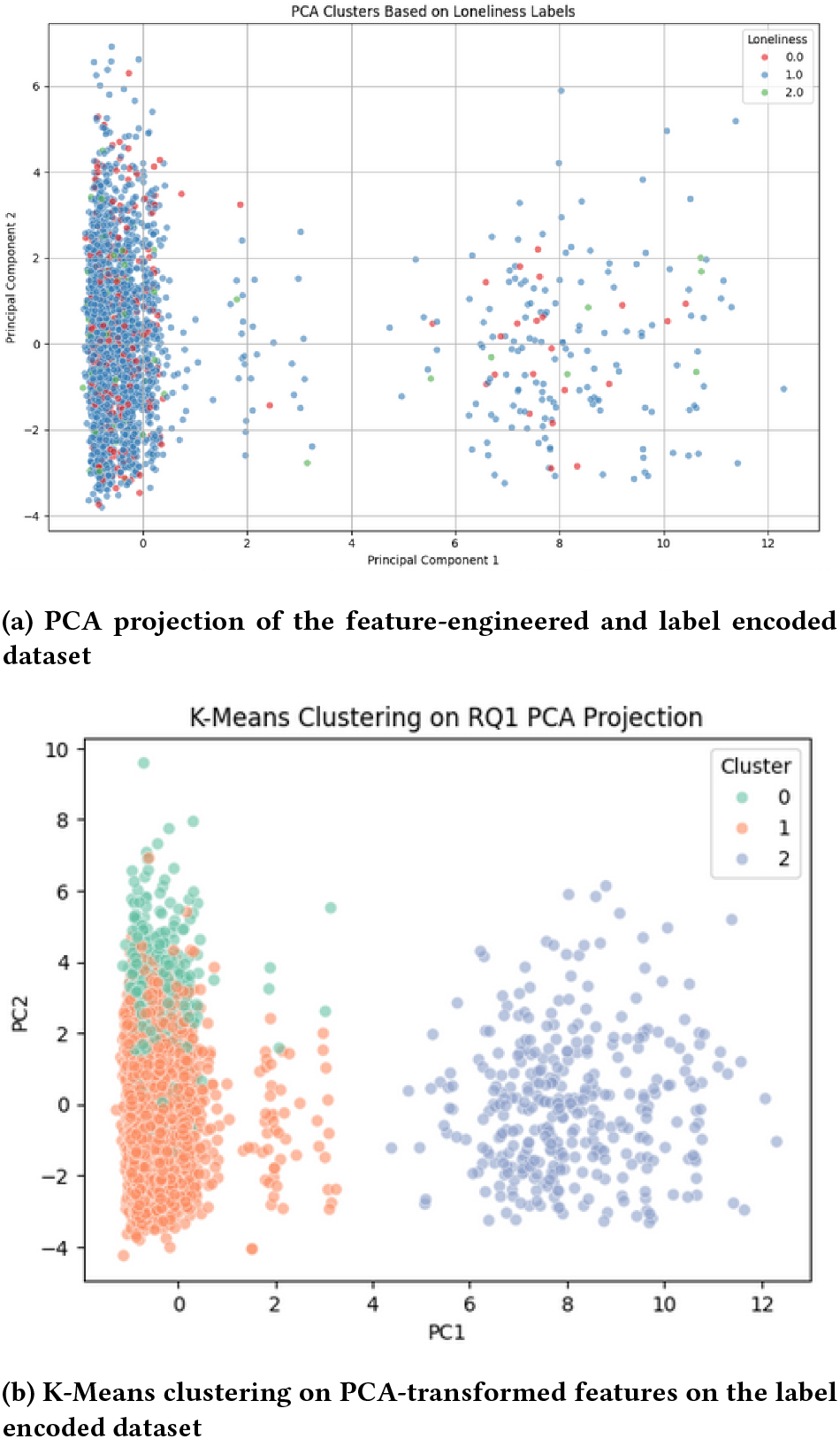
PCA and K-Means visualisations showing latent structure after feature engineering and label encoding.

Table 2 shows the transformed dataset, corresponding to the raw representation in Table 1. Target-encoded features also facilitated univariate feature-importance estimation, enabling selection of the top 50 predictors while retaining key behavioral and sociodemographic signals.

**Table 2:** Excerpt of the encoded feature values of the merged dataset.

|  | country_of_birth_encoded | exrta_domestic_current_assisted_living_encoded | activity_walking_pleasure_duration_encoded |
| --- | --- | --- | --- |
| 0 | 0.881188 | 0.918081 | 0.896552 |
| 1 | 0.921434 | 0.916101 | 0.906415 |
| 2 | 0.881579 | 0.916101 | 0.918877 |
| 3 | 0.913163 | 0.913014 | 0.921784 |
| 4 | 0.913163 | 0.913014 | 0.892308 |
| ⋮ | ⋮ | ⋮ | ⋮ |
| 5549 | 0.923539 | 0.918081 | 0.928571 |
| 5550 | 0.922928 | 0.920172 | 0.913303 |
| 5551 | 0.921434 | 0.916101 | 0.923469 |
| 5552 | 0.922928 | 0.920172 | 0.913303 |
| 5553 | 0.922928 | 0.920172 | 0.892857 |

### 3.3 Feature Selection

To derive a compact and interpretable subset of predictors, we applied Recursive Feature Elimination (RFE) using a logistic regression estimator to the 50-feature target-encoded matrix. RFE iteratively removed the least informative variables based on model-estimated coefficients, recursively refining the feature space to identify the most influential predictors of loneliness. Across repeated cross-validation folds, the procedure converged consistently on a stable subset of 15–20 features, indicating robustness of the selected attributes under resampling variability. This reduced feature set constitutes the foundation of the feature-engineered modelling pipeline. Based on the final RFE configuration, the selected features are as follows:

~~~
(1) health_mental_past_two_week_depression_frequency_encoded
(2) health_mental_current_happiness_encoded
(3) health_mental_current_satisfaction_friends_relations_encoded
(4) health_mental_current_satisfaction_family_relations_encoded
(5) country_of_birth_encoded
(6) activity_walking_pace_encoded
(7) activity_social_visit_frequency_encoded
(8) activity_spread_type_used_more_encoded
(9) diet_cereal_intake_choice_encoded
(10) diet_cheese_intake_encoded
(11) diet_milk_type_used_more_encoded
(12) alcohol_monthly_red_intake_choice_encoded
(13) sleep_nap_encoded
(14) sleep_daytime_encoded
~~~

The resulting feature subset captures core demographic, psychological, and behavioural determinants of psychological well-being. Mental-health indicators (Features 1–2) and relational-satisfaction measures (Features 3–4) represent direct psychological correlates of loneliness. Demographic context is reflected in country_of_birth_encoded (Feature 5), while behavioural activity patterns (Features 6–7) and lifestyle habits includes diet and alcohol consumption (Features 9–12) provide broader ecological context. Finally, sleep-related variables (Features 13–14) contribute physiological dimensions associated with mood regulation and social functioning. In combination, these features provide a concise representation of the multi factorial determinants associated with loneliness, directly addressing RQ1 and providing a foundation for subsequent analyses of class-associated configurations relevant to RQ2.

### 3.4 Machine-Learning Models

After preprocessing and feature selection, we constructed two supervised learning pipelines for comparison. In the first pipeline, the models were trained on the complete set of 50 target-encoded features. In the second pipeline, training was limited to the 15 predictors selected via RFE, as described in Section 3.3.

Logistic Regression, SVM, Random Forest, Gradient Boosting, CatBoost [19], and LightGBM [8] were trained under both pipelines to assess robustness across linear, kernel-based, and ensemble models. Models were trained using an 80/20 train-validation split to enable relative comparison across modeling configurations. Performance was evaluated using overall accuracy, class-specific accuracy, precision, recall, and F1-score. This dual-pipeline design enables direct comparison between models trained on the full feature set and a compact, domain-relevant subset.

To complement traditional tabular models, a reasoning-oriented large language model (LLM), `DeepSeek-R1-Distill-Qwen-1.5B`, was fine-tuned on text-based representations of participant attributes. The model was selected for its strong few-shot performance, computational efficiency, and demonstrated suitability for structured prediction tasks [23]. The LLM is not intended as a replacement for tabular classifiers, but as a probe for whether contextual reasoning over heterogeneous psychosocial attributes captures interaction-level signals that are difficult to model explicitly.

Participant records were reformatted into natural-language profiles summarizing psychological, social, behavioural, and demographic characteristics. Fine-tuning was conducted using a three-class sequence-classification objective (*Yes, No, Do not know*), with Parameter-Efficient Fine-Tuning (PEFT) [12] applied via the accelerate framework over three epochs. The model was evaluated on both (i) the target-encoded representation and (ii) the raw textual data, enabling comparison of encoding effects on LLM performance. A Chain-of-Thought (CoT) prompting strategy was also employed to elicit structured reasoning, with the prompt explicitly grounding predictions in observed feature values. To mitigate hallucinations, we restricted LLM inputs to only the 15 RFE-selected features, excluding raw text or unstructured data and applied temperature sampling (*T* = 0.3) to reduce randomness in responses. This ensures that LLM reasoning remains traceable to input data and reduces reliance on spurious patterns. The prompt below showcases an example of how it integrates mental health indicators, social connectedness, and life circumstances to guide prediction.

~~~
You are a clinical psychologist analyzing patient data to assess loneliness.
Patient Profile:
{features_block}
Task: Based on the patient’s mental health status, social connections, demographics, and other factors, predict whether they currently feel lonely.
Consider:
- How mental health symptoms (depression, anxiety) relate to loneliness
- The strength and quality of social connections
- Life circumstances that may contribute to or protect against loneliness
- Interactions between multiple factors
Answer with one of: Yes, No, Do not know.
~~~

This setup allows direct comparison between LLM-based contextual reasoning and conventional tabular classifiers, facilitating analysis of cases where predictions are driven by interactions among multiple attributes rather than isolated features.

### 3.5 Feature Contribution Analysis

To operationalize interpretability, we quantified feature contributions using univariate logistic regression augmentation, where each feature was incrementally added to a baseline model, and changes in accuracy, F1-score, coefficient magnitude, and odds ratios were recorded. This produced a ranked list of features by their predictive influence (Table 5), which we cross-referenced with psychological literature to validate alignment with known determinants of loneliness (e.g., social visit frequency [7], sleep patterns [21]). The analysis highlights the demographic, behavioral, and psychological attributes most strongly associated with loneliness risk within the HPP cohort and informs the subsequent results discussion.

## 4 Ethics Statement

This study primarily uses secondary, de-identified data from the Human Phenotype Project (HPP), accessed under the project’s data-use agreement. All participant data were pseudonymised numerically prior to access, and no directly identifying information was available to the authors. The original data collection procedures received approval from the appropriate institutional ethics committees, and all participants provided informed consent. The present analysis involved no additional data collection and was conducted in accordance with relevant ethical guidelines and regulations.

## 5 Results and Discussion

This section presents empirical findings from machine-learning models, large language models, and feature contribution analyses, illustrating how feature representation and selection impact loneliness classification within the HPP cohort. Interpretability is defined here as transparency in feature selection and model comparison, rather than causal inference or individual-level clinical explanation. To obtain a compact set of informative predictors, Recursive Feature Elimination (RFE) was applied to the 46 target-encoded features, identifying 15 that were consistently stable and discriminative across cross-validation folds. This subset balances predictive performance with interpretability and forms the basis of the feature-engineered modeling pipeline.

### 5.1 Machine Learning Model Performances

Table 3 reports classifier performance prior to feature engineering, while Table 4 presents results after feature engineering. Overall accuracy is driven by the dominant “Not Lonely” class, inflating metrics despite low Macro F1 scores. Ensemble methods like Random Forest and CatBoost achieve high accuracy but poor recall for minority classes, reflecting limited sensitivity to underrepresented states. Feature engineering improves minority-class detection across several classifiers. Logistic Regression shows increased Lonely recall (0.581− 0.601) without loss of weighted F1, while Gradient Boosting and LightGBM demonstrate modest gains in Lonely F1, suggesting that the refined feature set better captures loneliness-related patterns.

**Table 3:** Classifier performance before feature engineering.

| Classifier | Accuracy | Macro F1 | Weighted F1 | Lonely Recall | Lonely Precision | Lonely F1 | Not Lonely Recall | Don't Know Recall |
| --- | --- | --- | --- | --- | --- | --- | --- | --- |
| Logistic Regression | 0.665 | 0.455 | 0.730 | 0.601 | 0.347 | 0.440 | 0.684 | 0.407 |
| Random Forest | 0.861 | 0.417 | 0.821 | 0.217 | 0.642 | 0.325 | 0.988 | 0.000 |
| Gradient Boosting | 0.845 | 0.473 | 0.830 | 0.414 | 0.503 | 0.454 | 0.939 | 0.034 |
| SVM | 0.776 | 0.436 | 0.788 | 0.449 | 0.336 | 0.384 | 0.852 | 0.051 |
| Naive Bayes | 0.220 | 0.238 | 0.289 | 0.475 | 0.308 | 0.374 | 0.171 | 0.525 |
| LightGBM | 0.851 | 0.475 | 0.834 | 0.404 | 0.513 | 0.452 | 0.948 | 0.034 |
| CatBoost | 0.863 | 0.449 | 0.834 | 0.333 | 0.574 | 0.422 | 0.973 | 0.000 |

**Table 4:** Classifier performance after feature engineering.

| Classifier | Accuracy | Macro F1 | Weighted F1 | Lonely Recall | Lonely Precision | Lonely F1 | Not Lonely Recall | Don't Know Recall |
| --- | --- | --- | --- | --- | --- | --- | --- | --- |
| Logistic Regression | 0.699 | 0.462 | 0.760 | 0.581 | 0.382 | 0.461 | 0.734 | 0.271 |
| Random Forest | 0.861 | 0.441 | 0.830 | 0.303 | 0.577 | 0.397 | 0.976 | 0.000 |
| Gradient Boosting | 0.836 | 0.488 | 0.827 | 0.424 | 0.444 | 0.434 | 0.926 | 0.085 |
| SVM | 0.704 | 0.436 | 0.753 | 0.561 | 0.321 | 0.408 | 0.747 | 0.153 |
| Naive Bayes | 0.205 | 0.257 | 0.278 | 0.444 | 0.456 | 0.450 | 0.152 | 0.644 |
| LightGBM | 0.839 | 0.485 | 0.828 | 0.399 | 0.444 | 0.420 | 0.932 | 0.085 |
| CatBoost | 0.860 | 0.443 | 0.831 | 0.318 | 0.553 | 0.404 | 0.972 | 0.000 |

**Table 5:** Incremental feature addition results showing model performance as features are added sequentially.

| Num_Features | CV_Accuracy_Mean | CV_Std | CI_95_Lower | CI_95_Upper | Test_Accuracy |
| --- | --- | --- | --- | --- | --- |
| 1 | 0.8536 | 0.0014 | 0.8508 | 0.8564 | 0.8488 |
| 2 | 0.8626 | 0.0051 | 0.8526 | 0.8726 | 0.8512 |
| 3 | 0.8637 | 0.0095 | 0.8451 | 0.8822 | 0.8524 |
| 4 | 0.8660 | 0.0061 | 0.8541 | 0.8779 | 0.8578 |
| 5 | 0.8660 | 0.0056 | 0.8550 | 0.8769 | 0.8584 |
| 6 | 0.8665 | 0.0066 | 0.8536 | 0.8794 | 0.8596 |
| 7 | 0.8662 | 0.0070 | 0.8525 | 0.8799 | 0.8596 |
| 8 | 0.8655 | 0.0077 | 0.8504 | 0.8805 | 0.8638 |
| 9 | 0.8657 | 0.0078 | 0.8505 | 0.8810 | 0.8620 |
| 10 | 0.8670 | 0.0080 | 0.8514 | 0.8826 | 0.8692 |
| 11 | 0.8670 | 0.0071 | 0.8531 | 0.8808 | 0.8680 |
| 12 | 0.8675 | 0.0061 | 0.8555 | 0.8795 | 0.8692 |
| 13 | 0.8675 | 0.0061 | 0.8556 | 0.8794 | 0.8662 |
| 14 | 0.8675 | 0.0066 | 0.8547 | 0.8804 | 0.8668 |
| 15 | 0.8673 | 0.0069 | 0.8537 | 0.8808 | 0.8662 |

Tree-based classifiers maintain near-perfect recall for the “Not Lonely” class (0.926 − 0.988), consistent with the imbalanced class distribution, whereas performance on the “Do not know” class remains low across models, reflecting its sparse representation and potential label ambiguity. Naive Bayes performs poorly across all metrics, likely due to its strong conditional independence assumptions and sensitivity to skewed categorical distributions. Overall, feature engineering yields consistent yet modest improvements in minority-class recall and Macro F1 while preserving high accuracy, indicating enhanced sensitivity to loneliness-related patterns rather than mere redistribution of prediction errors.

### 5.2 Incremental Feature Analysis

To quantify the predictive influence of individual variables, we conducted an incremental feature analysis using Logistic Regression. Features from the 15 RFE-selected predictors (Section 3.3) were added sequentially, and model performance was recorded at each step (Table 5). The results in Table 5 show a steady increase in cross-validation accuracy as features are added, serving as our primary tool for operationalizing univariate feature importance. Each row represents the cumulative effect of adding one feature at a time, ranked by their contribution to predictive performance. This data-driven ranking directly addresses **RQ1** by quantifying how individual features (e.g., `activity_walking_pace_encoded`) influence loneliness prediction, aligning with psychological literature on physical activity and mood [21].

Incrementally adding features (Table 5) demonstrates how demographic, lifestyle, and social-health variables jointly predict loneliness, with the top five features (e.g., depression frequency, social visit frequency) having the strongest impact. Confidence intervals and standard deviations shrink as more features are included, highlighting loneliness as an interdependent, multi factorial outcome with no single dominant predictor.

Overall, this analysis confirms that loneliness emerges from the combined effect of multiple factors, justifying the selection of the final 15-feature set and motivating subsequent LLM-based classification.

### 5.3 LLM-Based Classification of Loneliness Labels

To complement traditional classifier performances, we evaluated the DeepSeek 1.5B LLM across four feature-processing configurations: (i) the full set of features after initial cleaning, (ii) a reduced set of 50 features derived via target encoding and mutual-information ranking, (iii) a refined set of 15 features selected via Recursive Feature Elimination (RFE), and (iv) their corresponding target-encoded variants. Table 6 summarizes the results.

**Table 6:**
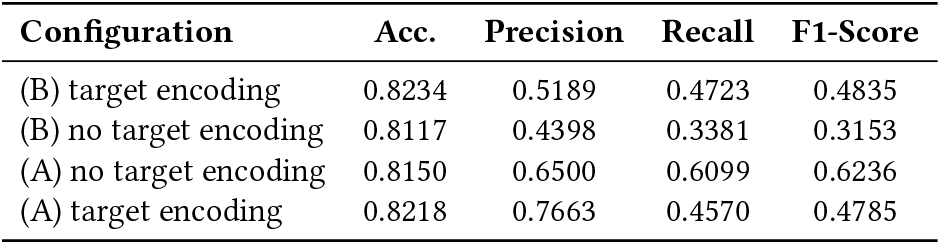
LLM performance across preprocessing setups. (B) = before feature engineering; (A) = after feature engineering with 15 features selected via RFE.

The strongest performance was achieved using the 15 feature-engineered predictors without target encoding, reaching 81% accuracy and the highest precision, recall, and F1-score. This suggests that while target encoding can reduce dimensionality for linear models, it may compress class-distinguishing semantic structures, limiting LLM performance. Feature-engineered data with target encoding maintained high precision but slightly lower accuracy, reflecting a trade-off between noise reduction and semantic richness. Despite CoT prompting, PEFT fine-tuning, and controlled feature selection, performance plateaued below 90%, indicating that capturing subtle psychosocial signals may require larger models, richer features, or hybrid tabular-LLM architectures.

The LLM-based classifier functions as an exploratory tool to assess whether high-level reasoning over structured feature interactions can uncover patterns missed by conventional models. CoT prompting aids interpretability but is not intended for clinical use, instead serving as a baseline for contextual reasoning in psychosocial classification.

#### Answer to RQ2

Loneliness reflects recurring combinations of psychological, social, and lifestyle factors rather than any single predictor. Across classical ML, feature-selection, and LLM analyses, improvements in minority-class detection emerge only when multiple domains are jointly considered, highlighting their interdependence. Consistent feature contributions across models reveal stable psychosocial patterns, supporting interpretable and scalable assessment approaches.

### 5.4 Impact and Implications

The results provide several insights for computational mental health monitoring and intervention design. Target encoding and feature selection enhances predictive performance and interpretability in complex psychosocial datasets. Through univariate feature analysis, we can identify key demographic, behavioural, and psychological predictors of loneliness, which supports the development of ethical and transparent screening tools. Moreover, using hybrid approaches on LLMs with tabular data demonstrates the potential for leveraging relational patterns in structured datasets, offering avenues for future model development. Collectively, these findings can inform population-level wellbeing initiatives by enabling early identification of individuals at risk of loneliness and guiding data-driven interventions.

## 6 Limitations and Ethical Considerations

This study is observational and cross-sectional, and it does not support causal inference between lifestyle, social, and demographic factors and psychological outcomes. The Human Phenotype Project dataset reflects specific demographic and cultural distributions, limiting generalizability. Class imbalance and resampling procedures may affect predictive performance and feature relevance, with imbalance-mitigation strategies potentially altering feature importance and reducing interpretability across subgroups; thus, findings should be interpreted at the population level rather than individually.

Ethically, inferred loneliness or psychological vulnerability should not be treated as diagnostic. Automated modeling carries risks of misclassification, stigmatization, and unintended consequences. LLM-based analyses in this study are strictly exploratory, intended for hypothesis generation and understanding feature interactions, not for individual-level assessment or clinical recommendation.

## 7 Conclusion

In conclusion, this study examines the multidimensional determinants of loneliness and psychological well-being using the HPP cohort, focusing on lifestyle, social, and demographic factors. Target encoding emerged as the most effective strategy for categorical variables, while PCA and clustering revealed meaningful latent structures. Feature engineering, particularly via Recursive Feature Elimination, improved model performance, especially for minority classes, and classical machine learning models such as Gradient Boosting and LightGBM robustly captured loneliness-related patterns. Incremental feature contribution analysis highlighted the multi factorial nature of loneliness, emphasizing the need for nuanced, data-driven approaches.

The integration of a Large Language Model (LLM) provided complementary insights into contextual and relational patterns. LLM performance was influenced by feature richness, with the best results achieved using a refined set of 15 features without target encoding, suggesting the value of preserving semantic diversity in numerical data. Overall, the findings demonstrate the importance of careful preprocessing, feature selection, and the combination of classical and modern machine learning approaches for modeling complex psychosocial constructs. These insights advance understanding of the interplay between lifestyle, social health, and psychological well-being, informing the development of data-driven interventions for loneliness and mental health support.

## Data Availability

All data referred to in this manuscript are available upon reasonable request through the Human Phenotype Project (https://knowledgebase.pheno.ai/platform_tutorial.html).

https://knowledgebase.pheno.ai

## Acknowledgments

We acknowledge the use of generative AI to assist with the language in polishing in this paper. All technical content, analysis, and conclusions are the authors’ own.

